# Safety & Effectiveness of Mifepristone for Medication Abortion in a National Claims Cohort, 2017-2023

**DOI:** 10.64898/2026.10.02.26364611

**Authors:** Liana R. Woskie, Julia Strasser, Risa Griffin, Qian (Eric) Luo, Maria I. Rodriguez, Ushma D. Upadhyay

**Affiliations:** Department of Community Health, Tufts University, Medford, Massachusetts, USA; Fitzhugh Mullan Institute for Health Workforce Equity, The George Washington University, Washington, DC, USA; Department of Obstetrics and Gynecology, Oregon Health & Science University, Portland, Oregon, USA; Department of Obstetrics, Gynecology, and Reproductive Sciences, University of California, San Francisco, San Francisco, California, USA

**Author notes:** **Corresponding author:** Ushma D. Upadhyay, PhD, MPH, Department of Obstetrics, Gynecology, and Reproductive Sciences, University of California, San Francisco, San Francisco, California, USA.

## Abstract

**BACKGROUND:** Extensive research has documented medication abortion safety. Large-scale insurance claims data offer an important complement to clinical studies by assessing real-world outcomes in routine practice. In claims data, researchers must infer mifepristone use, clinical outcomes, and event severity from diagnosis, treatment, and utilization codes. Understanding how these analytic choices influence safety and effectiveness estimates is important for interpreting evidence used to inform care delivery and medication policy.

**METHODS:** We conducted a retrospective cohort study of mifepristone use with national multipayer claims data, both medical and pharmacy, from 2017 through 2023. We identified both a broad cohort and a more specific medication-abortion cohort after excluding evidence of other indications. We classified potential safety events, effectiveness outcomes, and health care utilization within 45 days of mifepristone use. Serious adverse events required claims-observable consequences consistent with FDA seriousness criteria.

**RESULTS:** The broad cohort included 869,409 mifepristone uses among 698,780 unique patients for medication abortion or other indications. After restriction to uses with additional evidence of medication abortion, 775,931 uses among 626,443 unique patients remained. In the medication-abortion cohort, 3,306 uses were followed by at least one serious adverse event, resulting in an absolute 45-day risk of 0.43%. Serious bleeding or hemorrhage occurred after 0.24% of uses, serious infection or sepsis after 0.05%, serious hypersensitivity or anaphylaxis after 0.01%, and serious ectopic pregnancy outcomes after 0.05%.

**CONCLUSIONS:** Serious adverse events after mifepristone use were rare in this national study. Applying established, clinically grounded criteria markedly reduced the number of events classified as serious.

## INTRODUCTION

In 2000, the U.S. Food and Drug Administration (FDA) approved mifepristone for medication abortion, and it has since become widely used in the United States.^1^ More than one million abortions occur annually in the United States, and medication abortion accounts for the majority of clinician-provided abortions.^2^ Systematic reviews, large clinical cohorts, and population-based studies have consistently found serious adverse events after medication abortion to be rare.^3–6^ Despite a strong safety profile, mifepristone faces growing threats to its availability through proposed state and federal policy changes. Given these potential restrictions, estimates of the medication’s safety hold increasing clinical and regulatory importance.

Numerous studies, from clinical trials to observational studies using primary data collection or administrative databases, have substantiated the safety and effectiveness of in-person and telehealth medication abortion.^3–13^ A recent analysis of national insurance claims, however, reported markedly higher rates of serious adverse events after mifepristone use.^14^ Reconciling these differences is important because safety and effectiveness estimates inform both clinical care and regulation. Claims data have well-documented limitations for evaluating abortion; for example, many abortions are self-paid and may not appear in these data, and validated algorithms for reliably identifying abortion in claims remain limited.^15–17^ At the same time, administrative claims offer many advantages; they can capture care across large populations, allow for analysis of rare clinical events, have low attrition rates, allow for longitudinal following of patients regardless of health care provider or service, and are unlikely to be subject to reporting biases. Given their potential, what claims-based measures capture, and how analytic choices shape resulting estimates, warrants careful consideration.

These considerations are particularly relevant for mifepristone, which may also be captured in claims for indications or uses beyond medication abortion, including Cushing syndrome, pre-procedure preparation, or stillbirth. In claims-based studies, this creates several analytic decisions, including how to identify mifepristone use specific to first-trimester medication abortion, distinguish safety outcomes from effectiveness or follow-up care, and identify serious adverse events. Best practice in pharmacoepidemiologic research emphasizes that diagnosis or health care utilization codes may not, by themselves, establish the clinical nature or seriousness of an adverse event.^18,19^ FDA definitions distinguish serious adverse events based on consequences such as hospitalization, life-threatening events, disability, or medical or surgical intervention to prevent a serious outcome.^20,21^ Emergency department use, for example, reflects health care utilization rather than seriousness in itself. How foundational analytic choices, such as cohort construction and outcome classification, inform population-level safety estimates remains uncertain.

We therefore used a national multipayer health care claims database to characterize the 45-day risk of serious adverse events after mifepristone use from 2017 through 2023. Our primary empirical question was: How do estimates of post-mifepristone adverse events in insurance claims vary with cohort definition and application of clinical seriousness criteria? We first identified a broad cohort of mifepristone use and then a nested cohort with additional evidence of use for medication abortion. Within each cohort, we classified post-treatment events according to their primary clinical meaning and applied outcome-specific criteria for seriousness. We then compared estimates across cohorts and applied progressively more specific criteria to identify clinical events and assess their severity.

## METHODS

### Data Source and Study Population

We used deidentified longitudinal medical and pharmacy claims from the Komodo Health Healthcare Map from 2017 through 2023. The database includes commercial, Medicaid, and Medicare claims. Medical and pharmacy claims are linked longitudinally at the patient level and deduplicated across data sources. We identified mifepristone use using National Drug Codes and relevant procedure codes. Individuals could contribute more than one qualifying index treatment, or mifepristone use, during the study period. Each use was followed for 45 days to identify subsequent diagnoses, procedures, treatments, and health care utilization. Detailed code sets, and temporal rules for each step in our process are provided in the Supplementary Appendix. The George Washington University Institutional Review Board approved this study.

### Study Cohorts

We first constructed a broad mifepristone cohort using claims-based criteria designed to approximate the ascertainment approach described in a previous national claims analysis.^14^ Qualifying mifepristone uses were identified from mifepristone pharmacy or medical claims together with pregnancy-context information; for selected treatment codes, a corresponding diagnosis of encounter for elective termination of pregnancy (ICD-10-CM Z33.2) was also required. Diagnosis codes were not used alone to identify mifepristone use. Because the complete coding algorithm was not available in the previous national claims analysis,^14^ we selected diagnosis, procedure, treatment, and health care utilization codes to align as closely as possible with the published approach.

Because mifepristone is used for indications and clinical contexts other than medication abortion, we then constructed a nested medication-abortion cohort using a rule-out approach. We excluded cases where we had strong evidence of competing indications or treatment pathways, including Cushing syndrome, second-trimester or later pregnancy, fetal death or stillbirth established at or before index, planned procedural preparation, and clearly established preexisting spontaneous pregnancy loss. We retained mifepristone use with pregnancy-loss codes when other coding supported medication abortion.

### Outcome Classification

The primary outcome was total serious adverse events occurring within 45 days after mifepristone use. The analysis was a descriptive post-market safety characterization rather than a comparative safety analysis. We identified clinical events occurring within 45 days after each mifepristone use and classified them according to their primary clinical meaning before assessing severity. Potential safety events included hemorrhage, infection or sepsis, hypersensitivity or anaphylaxis, uterine or pelvic-organ injury, and other clinical events plausibly related to the abortion. We classified ectopic pregnancy separately as a pregnancy-location or diagnostic outcome. We classified additional mifepristone and/or misoprostol and subsequent procedural abortion as treatment-effectiveness outcomes according to the Medical Abortion Reporting of Efficacy framework.^22^ We evaluated health care utilization separately.

For selected outcome domains, we then applied progressively more specific criteria incorporating evidence of clinical evaluation, active treatment, and seriousness. These stages were outcome-specific and were intended to distinguish the initial diagnosis or utilization event from subsequent management and clinically serious outcomes.

### Seriousness Classification

We assessed seriousness using claims-observable consequences consistent with FDA criteria for serious adverse events.^20^ For prespecified clinical outcomes, a serious event generally required an underlying clinical event together with a qualifying consequence within the same encounter or contiguous clinical course. Qualifying criteria included blood transfusion, shock, major or urgent intervention, acute organ dysfunction, intensive or critical care, mechanical ventilation or clinically related inpatient hospital admission with active treatment. Emergency department visits or observation care alone did not establish seriousness.

We assigned seriousness criteria to the underlying clinical event when a corresponding diagnosis could be identified. To capture serious events that might otherwise be missed, we also included other serious clinical events plausibly related to the abortion that met qualifying seriousness criteria but did not fall within a prespecified outcome category. For ectopic pregnancy, we used a conservative definition that classified surgical management or another qualifying serious consequence as a serious ectopic outcome. Subsequent procedural abortion remained classified as an effectiveness outcome and contributed to the serious adverse event estimate only when accompanied by a qualifying serious clinical consequence.

### Statistical Analysis

We summarized characteristics of mifepristone use in the broad and nested cohorts using counts and percentages for categorical variables and means with standard deviations or medians with interquartile ranges for continuous variables, as appropriate. We calculated the Charlson comorbidity index using diagnoses observed during the 365 days before the index mifepristone use and categorized as 0 or ≥1 because of the low prevalence of comorbidity. For each clinical outcome, we estimated the 45-day absolute risk as the number of mifepristone uses with at least one qualifying event divided by the number of eligible mifepristone uses, with 95% confidence intervals.

To avoid counting a single event multiple times, we grouped related diagnoses, procedures, treatments, and care settings within the same acute clinical course. We counted each mifepristone use once for a given event or outcome. Because there could be more than one type of serious adverse event in a given post-treatment window, such as both hemorrhage and infection, we deduplicated the overall serious adverse event estimate at the treatment-episode level.

We estimated risk first in the broad mifepristone cohort and then in the nested medication-abortion cohort. We next examined how claims-based diagnoses and health care utilization narrowed with progressively more specific evidence of clinical evaluation, active treatment, and seriousness. We report component serious adverse events, treatment-effectiveness outcomes, and health care utilization separately, in addition to the deduplicated serious adverse event estimate.

## RESULTS

### Cohort Construction and Characteristics

We identified 869,409 mifepristone uses during the study period among 698,780 unique patients (Figure 1). After sequentially excluding mifepristone use with evidence of competing indications or treatment pathways, 775,931 mifepristone uses among 626,443 unique patients remained in the nested medication-abortion cohort. The nested cohort included 775,931 of 869,409 (89.2%) mifepristone uses in the broad mifepristone cohort, reflecting a relatively small reduction after application of the additional medication-abortion criteria (Figure 1).

**Figure 1.**
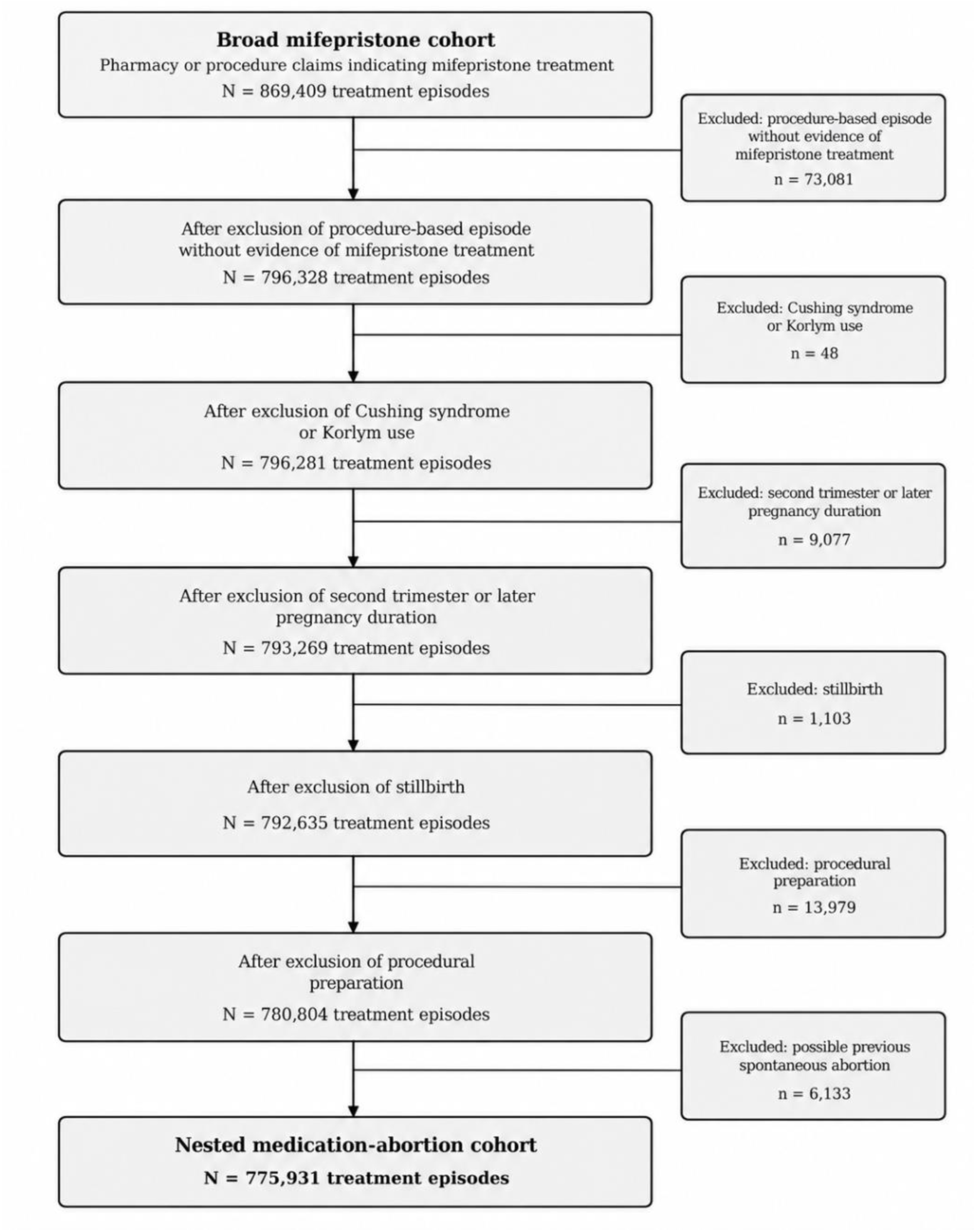
Construction of the Analytic Cohorts. **Note:** The broad mifepristone cohort included uses identified from mifepristone pharmacy or medical claims with pregnancy-context information; a diagnosis specifically identifying medication abortion at index was not required. The nested medication-abortion cohort used a sequential rule-out approach to increase specificity for early medication-abortion use while accommodating incomplete or discordant claims coding. Mifepristone uses were excluded for high-confidence evidence of competing indications or treatment pathways, including Cushing syndrome or Korlym use, second-trimester or later pregnancy, fetal death or stillbirth established at or before index, planned procedural preparation, or clearly established preexisting spontaneous pregnancy loss. Side-box counts show the total number of mifepristone uses meeting each exclusion criterion and are not mutually exclusive; they therefore may not equal the reduction between adjacent cohort counts. Pregnancy duration was based preferentially on the closest week-specific code on or before index, with trimester-specific codes used when week-specific information was unavailable. Post-index pregnancy-loss, procedural, hospitalization, and other acute-care claims were not used to redefine the index mifepristone use because they could reflect follow-up care, treatment effectiveness, or an adverse event. Ectopic pregnancies were retained for post-index outcome classification. An individual could contribute more than one qualifying mifepristone use. Detailed code sets and temporal rules are provided in the Supplementary Appendix.

Characteristics of the broad and nested cohorts are shown in Table 2. In the nested cohort, 1.6% of mifepristone uses occurred among persons 17 years of age or younger, 31.2% among those ages 18 to 24 years, 48.1% among those ages 25 to 34 years, and 16.8% among those ages 35 years or older. Across cohorts, Medicaid or Medicare was the most common payer for mifepristone use, accounting for 58.2% in the nested cohort, compared with 30.0% for commercial insurance. We were only able to estimate pregnancy duration for 14.0% of uses, and 93.4% of index mifepristone uses were identified through medical claims and 6.6% through pharmacy claims.

**Table 1.** Classification of Clinical Events and Seriousness following Mifepristone Use.

| Clinical event | Domain | Study classification and operational interpretation |
| --- | --- | --- |
| **Potential adverse events** |  |  |
| *Bleeding or hemorrhage* | Safety | For potential safety events, claims were first used to identify bleeding or hemorrhage, infection or sepsis, hypersensitivity or anaphylaxis, or uterine or pelvic-organ injury within 45 days after mifepristone use.<br>Seriousness was then assessed by identifying claims-observable consequences consistent with established Food and Drug Administration criteria for seriousness. A diagnosis alone generally did not establish seriousness unless it also involved a serious clinical event. |
| *Infection or sepsis* |  |  |
| *Hypersensitivity or anaphylaxis* |  |  |
| *Uterine or pelvic-organ injury* |  |  |
| *Other abortion-related clinical event* |  | Included to assess serious post-treatment events not captured by the specific outcome categories. Events were identified when claims supported a serious abortion-related clinical event meeting FDA seriousness criteria and were not already classified under another diagnosis listed above. |
| **Pregnancy-location and diagnostic events** |  |  |
| *Ectopic pregnancy* | Pregnancy location or diagnostic outcome | Ectopic pregnancy was not considered a treatment-associated adverse event by itself and did not necessarily indicate delayed diagnosis. Medical management with methotrexate alone was not considered serious. Surgical treatment of a confirmed ectopic pregnancy was included as a serious clinical outcome, as were ectopic pregnancies associated with shock, transfusion, sepsis, major rescue intervention, or another qualifying serious consequence. |
| **Effectiveness events** |  |  |
| *Additional mifepristone and/or misoprostol* | Effectiveness or follow up care | Classified according to the Medical Abortion Reporting of Efficacy (MARE) guidelines. Additional mifepristone or misoprostol within 45 days after index mifepristone use was not considered a serious adverse event by itself. If an effectiveness event occurred with a qualifying serious clinical event or consequence within the same acute clinical course, it was included in the overall serious adverse-event measure. |
| *Subsequent procedure* |  | Subsequent procedures, such as aspiration or dilation and curettage, within 45 days after index mifepristone use were classified as effectiveness or follow up care outcomes and were not considered serious adverse events by themselves. If a subsequent procedure occurred with a qualifying serious clinical event or consequence within the same acute clinical course, it contributed to the overall serious adverse-event measure. |
| **Health care utilization events** |  |  |
| *Abortion-related emergency department visit* | Utilization | Emergency department visits were treated as health care utilization, not as adverse events or evidence of seriousness on their own. Emergency department visits were considered abortion-related when an abortion-related diagnosis was observed within 2 days before or after the emergency visit. Visits associated with external causes of injury (such as, motor-vehicle collisions or assault) were excluded. Evaluation without a pathologic diagnosis or active treatment was considered observation care; serious events were classified according to qualifying seriousness codes. |

**Table 2.** Serious Adverse Events by Selected Patient and Treatment Characteristics within 45 Days of Mifepristone Use, Komodo Health Healthcare Map, 2017-2023.

| Characteristic | Broad Mifepristone Cohort |  |  | Nested Medication-Abortion Cohort |  |  |
| --- | --- | --- | --- | --- | --- | --- |
|  | Mifepristone Uses,<br>No. (%) | Serious<br>Adverse<br>Events, No. | 45-Day Risk,<br>% (95% CI) | Mifepristone Uses,<br>No. (%) | Serious<br>Adverse<br>Events, No. | 45-Day Risk,<br>% (95% CI) |
| **Overall serious adverse events** |  |  |  |  |  |  |
|  | 869,409 (100.0) | 3,930 | 0.45% (0.44%-0.47%) | 775,931 (100.0) | 3,306 | 0.43% (0.41%-0.44%) |
| **Age, yr** |  |  |  |  |  |  |
| ≤17 | 14,933 (1.7) | 41 | 0.27% (0.19%-0.36%) | 12,720 (1.6) | 33 | 0.26% (0.17%-0.35%) |
| 18–24 | 267,900 (30.8) | 1,111 | 0.41% (0.39%-0.44%) | 241,990 (31.2) | 973 | 0.40% (0.38%-0.43%) |
| 25–34 | 419,306 (48.2) | 2,013 | 0.48% (0.46%-0.50%) | 373,310 (48.1) | 1,705 | 0.46% (0.44%-0.48%) |
| ≥35 | 149,598 (17.2) | 739 | 0.49% (0.46%-0.53%) | 130,477 (16.8) | 575 | 0.44% (0.40%-0.48%) |
| Unknown | 17,672 (2.0) | 26 | 0.15% (0.09%-0.20%) | 17,434 (2.2) | 20 | 0.11% (0.06%-0.16%) |
| **Pregnancy duration\*** |  |  |  |  |  |  |
| 1<sup>st</sup> trimester |  |  |  |  |  |  |
| ≤7 wk | 83,708 (9.6) | 313 | 0.37% (0.33%-0.42%) | 81,032 (10.4) | 292 | 0.36% (0.32%-0.40%) |
| 8–10 wk | 26,929 (3.1) | 211 | 0.78% (0.68%-0.89%) | 24,944 (3.2) | 199 | 0.80% (0.69%-0.91%) |
| > 10–13 wk | 2,664 (0.3) | 30 | 1.13% (0.73%-1.53%) | 401 (<0.1) | 15 | 3.74% (1.88%-5.61%) |
| Wks unsp | 2,186 (0.3) | 12 | 0.55% (0.24%-0.86%) | 2,152 (0.3) | 12 | 0.56% (0.24%-0.87%) |
| 2<sup>nd</sup> trimester | 8,903 (1.0) | 131 | 1.47% (1.22%-1.72%) | 0 (0.0) | 0 | - |
| 3<sup>rd</sup> trimester | 174 (<0.1) | 4 | 2.30% (0.05%-4.55%) | 0 (0.0) | 0 | - |
| Not estimable | 744,845 (85.7) | 3,229 | 0.43% (0.42%-0.45%) | 667,402 (86.0) | 2,788 | 0.42% (0.40%-0.43%) |
| **Payer** |  |  |  |  |  |  |
| Medicaid/Medicare | 519,317 (59.7) | 2,670 | 0.51% (0.49%-0.53%) | 451,937 (58.2) | 2,241 | 0.50% (0.48%-0.52%) |
| Commercial | 253,677 (29.2) | 960 | 0.38% (0.35%-0.40%) | 232,498 (30.0) | 804 | 0.35% (0.32%-0.37%) |
| Other or unknown | 96,415 (11.1) | 300 | 0.31% (0.28%-0.35%) | 91,496 (11.8) | 261 | 0.29% (0.25%-0.32%) |
| **Index claim type** |  |  |  |  |  |  |
| Pharmacy claim | 52,455 (6.0) | 173 | 0.33% (0.28%-0.38%) | 51,372 (6.6) | 158 | 0.31% (0.26%-0.36%) |
| Medical claim | 816,954 (94.0) | 3,757 | 0.46% (0.45%-0.47%) | 724,559 (93.4) | 3,148 | 0.43% (0.42%-0.45%) |
| **Charlson comorbidity index†** |  |  |  |  |  |  |
| 0 | 842,800 (96.9) | 3,751 | 0.45% (0.43%-0.46%) | 751,228 (96.8) | 3,177 | 0.42% (0.41%-0.44%) |
| ≥1 | 26,609 (3.1) | 179 | 0.67% (0.57%-0.77%) | 24,703 (3.2) | 129 | 0.52% (0.43%-0.61%) |

**Note:** Serious adverse events were defined as the deduplicated total of events meeting the study's claims-observable criteria for seriousness within 45 days after index mifepristone use. The broad cohort used inclusive claims-based criteria for mifepristone use; the nested cohort applied additional criteria intended to identify uses more consistent with medication abortion. Risks are unadjusted and are presented descriptively. Pregnancy duration was estimated from diagnosis codes on the index mifepristone claim. Week-specific codes were used when available; trimester-specific codes were used when week-specific information was unavailable. The >10–13-week category reflects week-specific coding; the unspecified first-trimester category reflects trimester-specific coding without a week-specific code. Second- and third-trimester categories combine week-specific and trimester-specific codes. Mifepristone uses with insufficient information were classified as not estimable.
† The Charlson comorbidity index was calculated from diagnoses observed during the 365 days before index mifepristone use; scores were categorized as 0 or ≥1 because of the low prevalence of comorbidity in this population.

### Serious Adverse Events

Within 45 days after mifepristone use, 3,306 of 775,931 mifepristone uses in the nested medication-abortion cohort were followed by at least one serious adverse event, corresponding to an absolute risk of 0.43% (95% CI, 0.41 to 0.44). Risk varied modestly across age and payer groups, from 0.26% among persons 17 years of age or younger to 0.46% among those 25 to 34 years of age, and was 0.50% among Medicaid- or Medicare-covered mifepristone uses compared with 0.35% among commercially insured uses. Corresponding estimates were 0.31% for uses identified through a pharmacy claim and 0.43% for those identified through a medical claim. Among uses with estimable pregnancy duration, risk was 0.36% at 7 weeks or less and 0.80% at 8 to 10 weeks; estimates for later pregnancy duration were based on relatively few cases. Serious-event risk was 0.42% among uses with a Charlson comorbidity index of 0 and 0.52% among those with an index of 1 or greater.

### Clinical Classification of Post-Treatment Events

Broad claims-based diagnoses and health care utilization were substantially more frequent than events meeting criteria for seriousness (Figure 2). Among 26,681 uses with a bleeding or hemorrhage diagnosis, 23,961 included active diagnostic evaluation, 6,760 included active treatment, and 2,126 met criteria for a serious bleeding or hemorrhage event, representing 8.0% of uses with a bleeding or hemorrhage diagnosis and 0.24% of the broad cohort. Similarly, 2,182 mifepristone uses had an infection or sepsis diagnosis, 1,529 included active treatment, and 461 met criteria for serious infection or sepsis, representing 21.1% of uses with an infection or sepsis diagnosis and 0.05% of the broad cohort.

**Figure 2.**
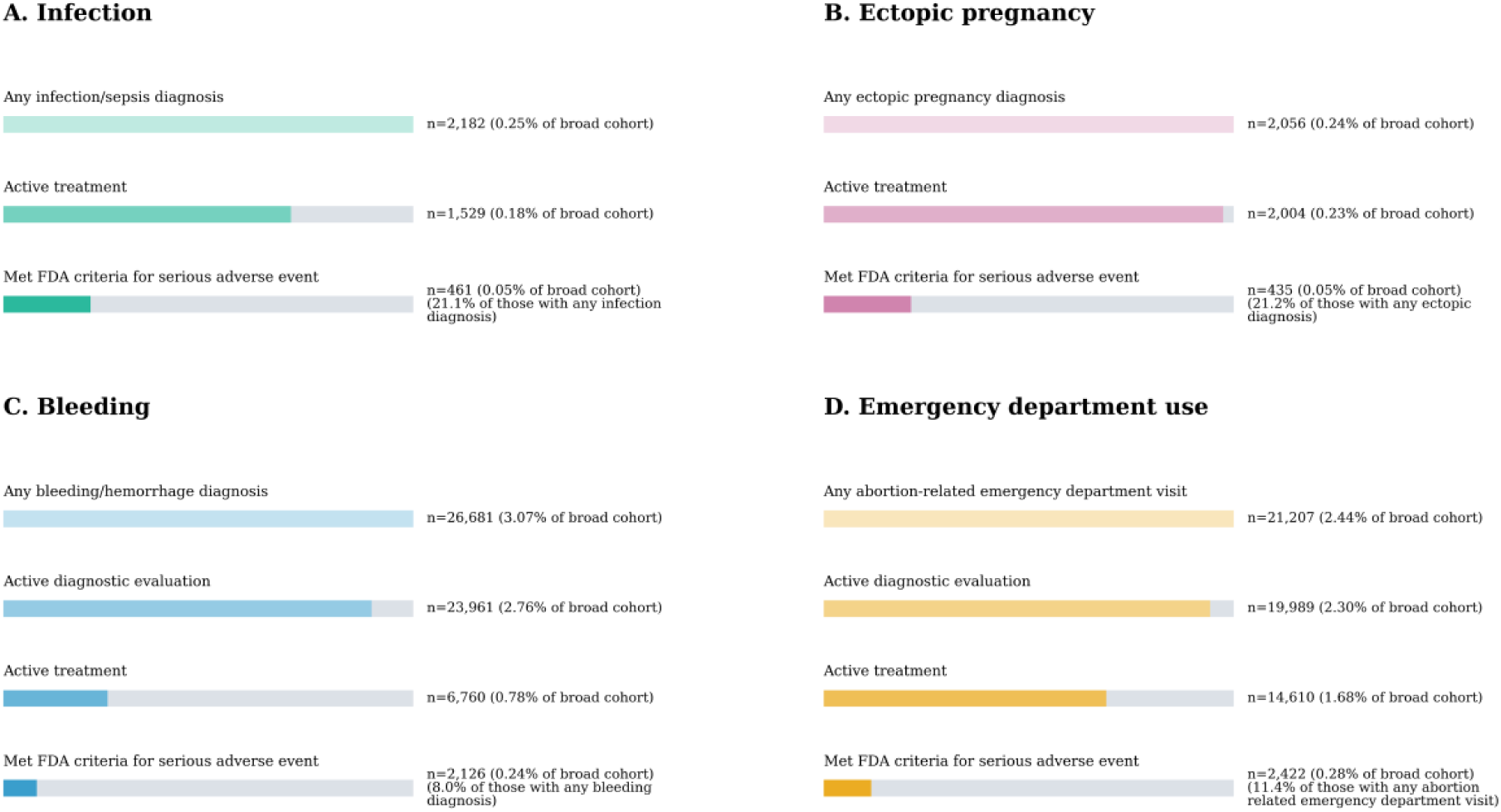
Clinical Cascades from Claims-Based Diagnoses and Utilization to Serious Adverse Events within 45 Days after Mifepristone Use, Komodo Health Healthcare Map, 2017–2023 **Note:** Each panel begins with a broad claims-based diagnosis or utilization event and applies progressively more selective criteria incorporating diagnostic evaluation, active treatment, and seriousness. Categories are cumulative within each panel, such that mifepristone uses meeting a later-stage criterion are also included in preceding stages. Active treatment indicates clinical management of the identified condition but does not, by itself, establish a serious adverse event. The final stage required a claims-observable consequence consistent with established FDA criteria for seriousness, such as: clinically related inpatient hospitalization with active treatment, transfusion, shock, major or urgent intervention, acute organ dysfunction, intensive or critical care, or mechanical ventilation. The emergency department panel represents a care setting rather than a distinct clinical diagnostic category and may therefore include uses also represented in the bleeding, infection, ectopic pregnancy, or other clinical-event categories. Not all serious adverse events identified in the study came through the emergency department; as such the emergency department SAE rate (0.28%) is lower than the study’s overall rate. The ectopic pregnancy panel is restricted to uses with a clear ectopic pregnancy diagnosis (ICD-10-CM O00.-). Bars are normalized to the initial claims signal within each panel. Annotations report the number of mifepristone uses and percentage of the broad mifepristone cohort (N=869,409); for the final stage in each panel, annotations additionally report the percentage of uses with the initial diagnosis or utilization event shown in that panel.

For ectopic pregnancy, 2,056 mifepristone uses had a qualifying diagnosis, 2,004 included active treatment, and 435 met criteria for a serious ectopic pregnancy outcome, representing 21.2% of uses with an ectopic pregnancy diagnosis and 0.05% of the broad cohort. An emergency department visit occurred after 66,695 uses; 21,207 were classified as abortion-related emergency department visits, 19,989 included active diagnostic evaluation, 14,610 included active treatment, and 2,422 were associated with a qualifying serious adverse event, representing 3.6% of uses with an emergency department visit and 0.28% of the broad cohort. Across domains, progressively incorporating evidence of clinical evaluation, treatment, and seriousness substantially narrowed the number of claims-defined events classified as serious adverse events.

### Events by Type

Among serious adverse events in the nested medication-abortion cohort, serious bleeding or hemorrhage was the most frequent, occurring after 1,872 mifepristone uses (0.24%) (Table 3). Serious infection or sepsis occurred after 372 uses (0.05%), serious hypersensitivity or anaphylaxis after 109 (0.01%), and serious uterine or pelvic-organ injury after 14 (<0.01%). Serious ectopic pregnancy outcomes, classified separately as pregnancy-location outcomes, occurred after 370 uses (0.05%). An additional 755 uses (0.10%) included a serious event captured in the other safety-related adverse-event category.

**Table 3.** Serious Adverse Events and Additional Post-Treatment Events within 45 Days of Mifepristone Use, Komodo Health Healthcare Map, 2017-2023.

| Outcome | Broad Mifepristone Cohort<br>(N = 869,409) |  | Nested Medication-Abortion Cohort<br>(N = 775,931) |  |
| --- | --- | --- | --- | --- |
|  | No. of Events | 45-Day Risk, %<br>(95% CI) | No. of Events | 45-Day Risk, %<br>(95% CI) |
| **Overall serious adverse events** |  |  |  |  |
| *Any serious adverse event\** | 3930 | 0.45%<br>(0.44%-0.47%) | 3306 | 0.43%<br>(0.41%-0.44%) |
| **Serious adverse events, safety** |  |  |  |  |
| *Serious bleeding or hemorrhage* | 2126 | 0.24%<br>(0.23%-0.25%) | 1872 | 0.24%<br>(0.23%-0.25%) |
| *Serious infection or sepsis* | 461 | 0.05%<br>(0.05%-0.06%) | 372 | 0.05%<br>(0.04%-0.05%) |
| *Serious hypersensitivity or anaphylaxis* | 133 | 0.02%<br>(0.01%-0.02%) | 109 | 0.01%<br>(0.01%-0.02%) |
| *Serious uterine or pelvic-organ injury* | 35 | 0.00%<br>(0.00%-0.01%) | 14 | 0.00%<br>(0.00%-0.00%) |
| *Other safety-related adverse events* | 981 | 0.11%<br>(0.11%-0.12%) | 755 | 0.10%<br>(0.09%-0.10%) |
| **Serious adverse events, pregnancy-location** |  |  |  |  |
| *Serious ectopic pregnancy outcome* | 435 | 0.05%<br>(0.05%-0.05%) | 370 | 0.05%<br>(0.04%-0.05%) |
| **Additional post-treatment events†** |  |  |  |  |
| **Effectiveness events‡** |  |  |  |  |
| *Additional mifepristone and/or misoprostol* | 24150 | 2.78%<br>(2.74%-2.81%) | 19732 | 2.54%<br>(2.51%-2.58%) |
| *Subsequent procedure* | 35079 | 4.03%<br>(3.99%-4.08%) | 18229 | 2.35%<br>(2.32%-2.38%) |
| **Health care utilization events** |  |  |  |  |
| *Abortion-related emergency department visit* | 21,207 | 2.44%<br>(2.41%-2.47%) | 19,363 | 2.50%<br>(2.46%-2.53%) |

\*A given mifepristone use could be associated with more than one serious adverse event, for example, both serious bleeding and serious infection following the same index use. The overall serious adverse event count was therefore deduplicated at the level of mifepristone use, whereas category-specific counts include all qualifying event types associated with each use and may sum to more than the overall count.
**Note:** Serious adverse events were identified using the classification and event-specific seriousness criteria summarized in Table 1. Seriousness was assessed using claims-observable consequences consistent with established Food and Drug Administration criteria. Related diagnosis, procedure, treatment, and care-setting claims occurring within the same acute clinical course were grouped to avoid counting a single event multiple times. Effectiveness events and emergency department visits are shown separately because they were not considered serious adverse events by themselves. Clinically related inpatient hospitalization with active treatment was a criterion for seriousness and therefore could contribute to the overall serious adverse-event total. If an effectiveness or utilization event occurred during the same clinical course as a qualifying serious event, the serious event was classified under the relevant serious adverse-event category.
† Effectiveness events and health care utilization events do not contribute to the overall serious adverse-event estimate. Additional medication, subsequent procedure, and emergency department use were not considered serious adverse events by themselves. Clinically related inpatient hospitalization with active treatment did meet the study criteria for seriousness.
‡ May include continuing/ongoing pregnancy

Post-treatment effectiveness events, which did not by themselves constitute serious adverse events, were more common. We identified additional mifepristone and/or misoprostol after 19,732 uses (2.54%), and a subsequent procedure after 18,229 (2.35%). These outcomes contributed to the serious adverse event measure only when accompanied by a qualifying serious clinical consequence. We identified abortion-related emergency department visits after 19,363 mifepristone uses (2.50%), which were evaluated separately as a measure of health care utilization.

## DISCUSSION

In this national claims-based study of mifepristone use, serious adverse events were rare, occurring after 0.43% of uses in the nested medication-abortion cohort. Claims-based diagnoses and health care encounters were substantially more frequent, but only a subset were accompanied by clinical evidence meeting criteria for seriousness. Refining the broad mifepristone cohort to uses more consistent with medication abortion had minimal effect on the overall serious adverse-event estimate, whereas estimates were substantially lower when diagnoses, treatment, and health care utilization were distinguished from events meeting established criteria for seriousness. These findings underscore the importance of incorporating clinical context and severity when administrative claims are used to characterize medication safety.

Claims data can be well suited for identifying rare serious outcomes that generate observable medical care, including hospital admission, transfusion, surgery, or other high-acuity treatment.^4,6^ A challenge is that diagnosis, treatment, and utilization codes do not themselves establish clinical severity. A bleeding diagnosis or uterine procedure may simply represent clinically relevant care. Prior research on emergency department visits following abortion has found utilization may involve observation without treatment; for example, in a national study of abortion-related emergency department visits, 51% involved observation only.^21^ Studies of early pregnancy care more broadly demonstrate that emergency departments commonly serve as sites of routine care.^9,21,29,30^ Greater distance from an abortion provider has also been associated with greater use of emergency departments, suggesting care location may reflect the access environment.^31^ Incorporating treatment context and claims-observable consequences can therefore help distinguish expected symptoms, routine or nonserious management, and health care utilization from clinically serious events.

Accurate identification of treatment context is important due to mifepristone’s use beyond medication abortion, including for early pregnancy loss, cervical preparation, and management of fetal death.^23–25^ We anticipated that misclassification of use could increase claims-based adverse-event estimates. However, restricting the broad cohort to presumed use for medication abortion produced minimal change, and the serious adverse-event estimate remained below 0.50%. This consistency is notable given concerns about abortion ascertainment in claims. Subsequent procedures may reflect treatment completion, ongoing or incomplete abortion, retained tissue, patient preference, or management of a complication rather than a serious adverse event.^3,5^ Bleeding is similarly susceptible to overclassification because it is expected with medication abortion, whereas serious hemorrhage is better identified by consequences such as transfusion, hospitalization, shock, or major intervention.^4,5^

Ectopic pregnancy illustrates the limits of treating all post-treatment diagnoses or procedures as adverse drug events. Medication abortion may be initiated in selected patients before an intrauterine pregnancy has been confirmed, with subsequent follow-up used to establish pregnancy resolution or identify an ectopic pregnancy.^26,27^ An ectopic pregnancy identified after mifepristone may therefore reflect a pregnancy whose location was unresolved when treatment began, and surgical treatment can represent appropriate management rather than evidence of drug toxicity. We conservatively classified surgically managed ectopic pregnancy as a serious post-treatment outcome, while recognizing that this approach may overstate the proportion representing treatment-related serious adverse events.

This study has several limitations. The estimates are descriptive rates of outcomes observed after mifepristone use and should not be interpreted as causal effects of the drug. Claims are generated for billing rather than research and may contain incomplete or inaccurate diagnostic, procedural, and treatment information, with limited clinical detail available to adjudicate individual events.^15–17^ Pregnancy duration could not be reliably determined for most mifepristone use. In addition, the study captures mifepristone use and subsequent care that generated insurance claims. Abortion is frequently paid for outside insurance, including among patients with coverage, and medication abortion provided through telehealth or other pathways outside conventional insurance billing may therefore be underrepresented.^28^ However, this limitation primarily affects which abortions are represented rather than outcome ascertainment within the cohort. If patients whose abortion care is captured through insurance have greater clinical complexity or health care use, this form of selection would make our estimates conservatively high.

In this large national cohort, serious adverse events after mifepristone use were rare and consistent with a broad evidence base on medication-abortion safety.^3–6^ Applying established, clinically grounded criteria helped distinguish serious outcomes from broader post-treatment diagnoses, treatment, and health care utilization. More broadly, our findings illustrate the importance of transparently specifying how cohort and outcome definitions shape claims-based safety estimates used in clinical and regulatory decision-making.

## Supporting information

Supplement, Code List

## Data Availability

Data availability: The data used in this study were obtained from the Komodo Health Healthcare Map and are proprietary; therefore, the underlying individual-level data cannot be made publicly available by the authors. Analytic code and code lists used to define study cohorts and outcomes will be made publicly available.

