## Supplement, Code List for "Safety & Effectiveness of Mifepristone for Medication Abortion in a National Claims Cohort, 2017-2023"

**Appendix Table A. Broad Cohort Inclusion**

| ***Code*** | ***Additional requirement(s)*** |
| --- | --- |
| S0199 |  |
| S0190 | Z33.2 |
| Mifepristone NDCs: 00591-4390-96, 0591-4390-35, 43393-0001-01, 43393-0001-06, 64875-0001-01, 64875-0001-03, 76346-0073-01, 76346-0073-02, 76346-0654-03 |  |
| S0190/ S0191/ J3490 | Z33.2 |
| J8499 | Z33.2 + patient state = CT, NJ, NM, NY, NC, PA, VT |
| X7724/ X7726/ Z0336 | Z33.2 + patient state = CA |
| S01091 | Z33.2 + patient state = GA |
| W7576/ W7577/ W7578/ W7579 | Z33.2 + patient state = IL |
| W0486 | Z33.2 + patient state = NJ |
| J4999/ 99070 | Z33.2 + patient state = NM |
| W0846/ 4849A/ 59899/ 58999 | Z33.2 + patient state = NY |

**Appendix Table B. Nested Cohort Exclusions**

| ***Exclusion category*** | ***Exclusion Codes*** | ***Additional requirement(s)*** | ***Timing relative to mifepristone use*** |
| --- | --- | --- | --- |
| Other procedure, no evidence of mifepristone provided | State-specific code + Z33.2, but no clear evidence of mifepristone use for medication abortion (S0199 code, S0190+Z332, 200mg mifepristone NDC code (43393-0001-01, 43393-0001-06, 64875-0001-01, 64875-0001-03)) |  | Day of index mifepristone dose |
| Cushings Disease/ Korlym | NDC: 00591-4390-96, 0591-4390-35, 76346-0073-01, 76346-0073-02, 76346-0654-03  ICD10CM: E24.0, E24.1, E24.2, E24.3, E24.4, E24.8, E24.9 | NO S0199 or mifepristone NDC (43393-0001-01, 43393-0001-06, 64875-0001-01, 64875-0001-03) | Day of index mifepristone dose |
| Second trimester or later pregnancy duration | Z3A.14, Z3A.15, Z3A.16, Z3A.17, Z3A.18, Z3A.19, Z3A.2, Z3A.3, Z3A.4, Z34.82, Z34.92, Z34.83. Z34.93, O09.92, O09.93 |  | Day of index mifepristone dose |
| Stillbirth | O36.4, O31.0, Z37.1, Z37.3, Z37.4, Z37.6, Z37.7, P95 |  | Day of index mifepristone dose – up to 3 days post index mifepristone dose |
| Procedural Prep | 59840, 59841 | location is not inpatient (inpatient claim or non-inpatient claim with place of service code 21 or 23) | Day of index mifepristone dose – up to 3 days post index mifepristone dose |
| Possible previous spontaneous abortion | O02.1, O03 |  | 30 days pre index mife dose – day of index mifepristone dose |

**Appendix Table C. Clinical Cascades (day after index mifepristone – 45 days post index mifepristone)**

| ***Clinical Cascade*** | ***Claims Codes*** | ***Additional Requirements*** |
| --- | --- | --- |
| Infection |  |  |
| Infection Diagnosis | O04.5, O03.0, O03.5, N71.0, N73.0, N73.2, N73.3, N73.5, N73.9, O048.7, O073.7 |  |
| Infection Active Treatment | J0530, J1271, J1230, H0020, S0030, J1836, J0696, J0735, J0694, J0456, J1953, J1100, J8540, S0077, J0736, J0737, 96365, 96366, 96367, 96368, 96374,  Infection Serious Adverse Event | Infection Diagnosis |
| Infection Serious Adverse Event | A4.0, A4.1, R65.20, R65.21, R57.2, 99291, intensive care revenue codes (0200, 0201, 0202, 0203, 0206, 0209), 5A1935Zx, 5A1945Zx, 5A1955Zx, O048.7, O073.7, hospitalization (bill type code = 011x OR (bill type code = 012x AND revenue code = 010x-021x) AND ICD10CM includes O/Z AND admit_type_code IS NOT 3, 4) | Infection Diagnosis |
| Ectopic Pregnancy |  |  |
| Ectopic Pregnancy Diagnosis | O00x |  |
| Ectopic Pregnancy Active Treatment | J9260, J9250, methotrexate NDCs (00703-3671-01, 00703-3675-01, 00703-3678-01, 16729-0277-03, 16729-0277-30, 16729-0277-35, 00143-9367-01, 00143-9830-01, 62332-0711-01, 62332-0711-05, 62332-0712-01, 46708-0711-01, 46708-0711-05, 46708-0712-01, 55150-0510-01, 55150-0510-05, 55150-0511-01, 55150-0511-10, 55150-0512-01, 55150-0512-10, 55150-0513-01, 61703-0350-37, 61703-0350-38, 61703-0408-41, 61703-0350-09, 61703-0350-10, 61703-0408-25, 61703-0124-40, 61703-0161-02, 61703-0161-05, 63323-0122-50, 63323-0123-10, 00143-9516-01, 00143-9517-01, 00143-9518-01, 00143-9519-10, 67457-0480-40, 16729-0516-11), 96372, 96401, 59120, 59121, 59130, 59135, 59136, 59140, 59150, 59151, 10T20ZZx, 10T23ZZx, 10T24ZZx, 10T27ZZx, 10T28ZZx, 59120, 59121, 59130, 59135, 59136, 59140, 59150, 59151, 59525, 0UT9x, 37244, 59120, 59121, 59130, 59135, 59136, 59140, 59150, 59151, 10T20ZZx, 10T23ZZx, 10T24ZZx, 10T27ZZx, 10T28ZZx,  Ectopic Pregnancy Serious Adverse Events | Ectopic Pregnancy Diagnosis |
| Ectopic Pregnancy Serious Adverse Event | O08.1, O08.2, O08.3, O08.4, O08.81, O08.82, R57.1, 36430, P9010, P9011, P9012, P9016, P9017, P9019, P9021, P9022, P9023, P9031, P9032, P9033, P9034, P9035, P9036, P9037, P9038, P9039, P9040, P9044, P9051, P9052, P9053, P9054, P9055, P9056, P9057, P9058, P9059, P9060, 302301, 302331, 302401, 302431, 5A1935Zx, 5A1945Zx, 5A1955Zx, 0UQ9x, 58150, 58152, 58180, 58200, 58210, 58260, 58262, 58263, 58267, 58270, 58275, 58280, 58285, 58290, 58291, 58292, 58293, 58294, 58541, 58542, 58543, 58544, 58548, 58550, 58552, 58553, 58554, 58570, 58571, 58572, 58573, 59120, 59121, 59130, 59135, 59136, 59140, 59150, 59151, 59525, 0UT9x, 37244, 59120, 59121, 59130, 59135, 59136, 59140, 59150, 59151, 10T20ZZx, 10T23ZZx, 10T24ZZx, 10T27ZZx, 10T28ZZx, hospitalization (bill type code = 011x OR (bill type code = 012x AND revenue code = 010x-021x) AND ICD10CM includes O/Z AND admit type code IS NOT 3 OR 4) | Ectopic Pregnancy Diagnosis |
| Bleeding |  |  |
| Bleeding Diagnosis | O04.6, O07.1, O03.1, O03.6, N93.8,  N93.9 | No non-abortion related trauma code (see External Injury Exclusions) |
| Bleeding Diagnostic Evaluation | 76817, 76830, 76856, 76857, 85025,  85027, 85014, 85018, 84702, 84703  Bleeding Active Treatment  Hemorrhage Serious Adverse Event | Bleeding Diagnosis |
| Bleeding Active Treatment | 59812, 58120,  Methylergonovine maleate, oral (0093-3655-22, 0093-3655-28, 69238-1605-2, 69238-1605-8, 27437-050-01, 27437-050-19, 27437-050-56, 27437-050-57, 60687-813-11, 60687-813-94, 0904-7282-10),  Methylergonovine maleate, injection (J2210, 0517-0740-01, 0517-0740-20, 51991-144-17, 63704-004-01, 51662-1556-1, 51662-1556-2, 51662-1668-1, 72162-1592-2, 63629-2074-1),  Oxytocin injection (J2590, 63323-012-03, 63323-012-06, 63323-012-02, 63323-012-11, 63323-012-10, 63323-012-30, 65219-021-00, 65219-021-01, 0143-9742-01, 0641-6114-01, 0641-6115-01, 55154-9584-5, 70518-4233-00, 70518-4233-01),  Carboprost tromethamine injection (0009-0856-05, 0009-0856-08, 0143-9442-01, 0143-9442-10, 0409-0064-10, 25021-477-01, 42571-435-54, 42571-435-72, 43598-179-10, 43598-179-11, 43598-698-11, 43598-698-58, 46708-777-01, 46708-777-10, 62332-777-01, 62332-777-10, 62559-900-01, 62559-900-11, 65145-132-01, 65145-132-10, 65219-579-00, 65219-579-01, 68083-584-10, 69784-240-01, 69784-240-10, 70121-1680-1, 70121-1680-7, 70512-859-01, 70512-859-05, 71839-137-01, 71839-137-10, 81665-202-01, 81665-202-10)  Tranexamic acid injection (0517-0960-01, 0517-0960-10, 14593-318-01, 14789-115-05, 14789-115-08, 23155-166-31, 23155-166-41, 25021-415-10, 25021-415-66, 25021-418-82, 43066-008-01, 43066-008-10, 60505-6169-0, 60505-6169-1, 63323-563-01, 63323-563-10, 65145-106-01, 65145-106-10, 67457-197-00, 67457-197-10, 72603-387-01, 72603-387-10, 81284-611-00, 81284-611-10, 81284-612-00, 81284-612-10, 83634-401-10, 83634-401-41),  Hemorrhage Serious Adverse Event | Bleeding Diagnosis |
| Bleeding Serious Adverse Event | O048.1, O073.1, R57.1, 36430 P9010 P9011 P9012 P9016 P9017 P9019 P9021 P9022 P9023 P9031 P9032 P9033 P9034 P9035 P9036 P9037 P9038 P9039 P9040 P9044 P9051 P9052 P9053 P9054 P9055 P9056 P9057 P9058 P9059 P9060 302301 302331 302401 302431 99291 99292 37244 58150 58152 58180 58200 58210 58260 58262 58263 58267 58270 58275 58280 58285 58290 58291 58292 58293 58294 58541 58542 58543 58544 58548 58550 58552 58553 58554 58570 58571 58572 58573 59525 0W3R0ZZx, 0W3R3ZZx, 0W3R4ZZx, 0W3R7ZZx, 0W3R8ZZx, 04LE0CZx, 04LE0ZZx, 04LE3CZx, 04LE3ZZx, 04LE4CZx, 04LE4ZZx, 04LF0CZx, 04LF0ZZx, 04LF3CZx, 04LF3ZZx, 04LF4CZx, 04LF4ZZx, 0UT9x, hospitalization (bill type code = 011x OR (bill type code = 012x AND revenue code = 010x-021x) AND ICD10CM includes O/Z AND admit type code IS NOT 3 OR 4) | Bleeding Diagnosis |
| Emergency Department Use |  |  |
| Any Abortion-Related Emergency Department (ED) Visit | 99281, 99282, 99283, 99284, 99285, Emergency Room revenue code (0450, 0451, 0452, 0456, 0459, 0981), Emergency Room place of service code (23) | Z332, Z640, O03, O04, O07, O072, O0731, O0732, O0736, O0737 within 2 days before or after ED visit;  No non-abortion related trauma code within 2 days before or after ED visit (see External Injury Exclusions) |
| Active Diagnostic Evaluation | 76817, 76830, 76856, 76857, 84702, 84703, 85025, 85027, 85014, 85018, ultrasound and laboratory testing revenue codes (0402, 030),  Active treatment,  ED Serious Adverse Event | ED Visit |
| Active Treatment | J2210, J2590, 59812, 58120, S0030, J1836, J0696, J0694, J0456, J0121, J3490, J3590, J0736, J0737, S0078, S0190, S0191, S0199, 59840, 59841, 59812, 59820, 59821, 58120, J9260, J9250, 96365, 96366, 96367, 96368, 96374, 96372, J1885, J1170, J2270, J3010, J2405, J2765, J2550, J0780, 96360, 96361, J7030, J7040, J7042, J7050, J7060, J7070, J7120, 96375, 96376, 16571-0735-17, 16571-0735-21, 16571-0735-28, 16571-0735-01, 70010-0786-12, 09047-2821-00, 27437-0050-57, 00093-3655-22, 00054-0639-13, 51991-0144-17, 00078-0054-61, 63323-0012-11, 63323-0012-03, 63323-0012-10, 63323-0012-30, 00641-6114-25, 00641-6114-01, 00641-6115-25, 65219-0021-01, 65219-0021-00, 00591-3553-69, 00009-0853-01, 00009-0853-02, 43598-0919-11, 43598-0919-58, 62332-0777-01, 62332-0777-10, 55150-0459-10, 71839-0137-01, 71839-0137-10, 81665-0202-01, 81665-0202-10, 05170-9600-01, 05170-9600-10, 14593-0318-01, 14789-0115-05, 14789-0115-08, 23155-0166-31, 23155-0166-41, 25021-0415-10, 25021-0415-66, 25021-0418-82, 43066-0008-01, 43066-0008-10, 60505-6169-00, 60505-6169-01, 63323-0563-01, 63323-0563-10, 65145-0106-01, 65145-0106-10, 67457-0197-00, 67457-0197-10, 72603-0387-01, 72603-0387-10, 81284-0611-00, 81284-0611-10, 81284-0612-00, 81284-0612-10, 83634-0401-10, 83634-0401-41, 00069-1005-01, 00143-9803-10, 60505-0018-00, 00338-1055-48, 00409-0152-24, 00093-0022-01, 00781-3206-95, 60505-6138-04, 00409-7332-01, 60505-6136-04, 60505-6137-04, 00143-9878-10, 00781-3072-95, 00093-7169-56, 00069-3060-75, 00409-6981-01, 00409-6982-02, 25021-0011-52, 00009-0331-02, 00703-3671-01, 00703-3675-01, 00703-3678-01, 16729-0277-03, 16729-0277-30, 16729-0277-35, 00143-9367-01, 00143-9830-01, 62332-0711-01, 62332-0711-05, 62332-0712-01, 46708-0711-01, 46708-0711-05, 46708-0712-01, 55150-0510-01, 55150-0510-05, 55150-0511-01, 55150-0511-10, 55150-0512-01, 55150-0512-10, 55150-0513-01, 61703-0350-37, 61703-0350-38, 61703-0408-41, 61703-0350-09, 61703-0350-10, 61703-0408-25, 61703-0124-40, 61703-0161-02, 61703-0161-05, 63323-0122-50, 63323-0123-10, 43393-0001-01, 43393-0001-06, 64875-0001-01, 64875-0001-03,  ED Serious Adverse Event | ED Visit |
| Serious Adverse Event | Serious Adverse Event – any clinical category | ED Visit |

**Appendix Table C1. External Injury Exclusions – excluded from bleeding diagnosis and emergency department visits**

| ***External Injury Exclusion category*** | ***Codes*** |
| --- | --- |
| Transport accidents | V00–V99 |
| Firearm injuries | W32–W34, X72–X74, X93–X95, Y22–Y24 |
| Assault-related stabbing or blunt trauma | X92–Y09 |
| Falls or other external accidents | W00–W19 |
| Major burns | T20–T32 |

**Appendix Table D. Additional Serious Adverse Events (not included in Clinical Cascades)**

| ***Clinical Event*** | ***Claims Codes*** | ***Additional Requirements*** | ***Timing relative to mifepristone use*** |
| --- | --- | --- | --- |
| Serious hypersensitivity or anaphylaxis | J0171, J0172, R579, 99291, 020.0, 020.1, 020.2, 020.3, 020.6, 020.9, 31500, 5A1935Z, 5A1945Z, 5A1955Z, T886A, T782A, hospitalization (bill type code = 011x OR (bill type code = 012x AND revenue code = 010x-021x) AND ICD10CM includes O/Z AND admit type code IS NOT 3 OR 4) | Hypersensitivity diagnosis: T783A, T7840A, T7849A, L500, T886A, T782A | Index mifepristone – 45 days post index mifepristone |
| Serious uterine or pelvic-organ injury | 0UQ9x, 37244, 58150, 58152, 58180, 58200, 58210, 58260, 58262, 58263, 58267, 58270, 58275, 58280, 58285, 58290, 58291, 58292, 58293, 58294, 58541, 58542, 58543, 58544, 58548, 58550, 58552, 58553, 58554, 58570, 58571, 58572, 58573, 59525, 0UT9x, 0UB9x, 36430, P9010, P9011, P9012, P9016, P9017, P9019, P9021, P9022, P9023, P9031, P9032, P9033, P9034, P9035, P9036, P9037, P9038, P9039, P9040, P9044, P9051, P9052, P9053, P9054, P9055, P9056, P9057, P9058, P9059, P9060 302301, 302331, 302401, 302431, O048.1, O073.1, R57.1, 99291, intensive care revenue codes (0200, 0201, 0202, 0203, 0206, 0209), 5A1935Zx, 5A1945Zx, 5A1955Zx, 44602x, 44603x, 44604x, hospitalization (bill type code = 011x OR (bill type code = 012x AND revenue code = 010x-021x) AND ICD10CM includes O/Z AND admit type code IS NOT 3 OR 4) | Uterine or pelvic-organ injury diagnosis: S3769A, O048.4, O073.4 | Day after index mifepristone – 45 days post index mifepristone |
| Other safety-related adverse events | O048.1, O073.1, R57.1, 36430, P9010, P9011, P9012, P9016, P9017, P9019, P9021, P9022, P9023, P9031, P9032, P9033, P9034, P9035, P9036, P9037, P9038, P9039, P9040, P9044, P9051, P9052, P9053, P9054, P9055, P9056, P9057, P9058, P9059, P9060, 302301, 302331, 302401, 302431, 99291, 37244, 58150, 58152, 58180, 58200, 58210, 58260, 58262, 58263, 58267, 58270, 58275, 58280, 58285, 58290, 58291, 58292, 58293, 58294, 58541, 58542, 58543, 58544, 58548, 58550, 58552, 58553, 58554, 58570, 58571, 58572, 58573, 59525, 0W3R0ZZx, 0W3R3ZZx, 0W3R4ZZx, 0W3R7ZZx, 0W3R8ZZx, 04LE0CZx, 04LE0ZZx, 04LE3CZx, 04LE3ZZx, 04LE4CZ 04LE4ZZx, 04LF0CZx, 04LF0ZZx, 04LF3CZ 04LF3ZZx, 04LF4CZx, 04LF4ZZx, A40x, A41x, R652.0, R652.1, R572x, intensive care revenue codes (0200, 0201, 0202, 0203, 0206, 0209), 5A1935Zx, 5A1945Zx, 5A1955Zx, 0UQ9x, 0UT9x, 0UB9x, hospitalization (bill type code = 011x OR (bill type code = 012x AND revenue code = 010x-021x) AND ICD10CM includes O/Z AND admit type code IS NOT 3 OR 4), O072x, O073.1, O073.2, O073.6, O073.7 | Abortion-related diagnosis code: Z33.2, Z64.0, O03x, O04x, O07x | Day after index mifepristone – 45 days post index mifepristone |

**Appendix Table E. Effectiveness Events**

| ***Clinical Event*** | ***Claims Codes*** | ***Timing relative to mifepristone use*** |
| --- | --- | --- |
| Additional mifepristone and/or misoprostol | S0190, S0191, S0199, mifepristone NDC (43393-0001-01, 43393-0001-06, 64875-0001-01, 64875-0001-03), misoprostol NDC (00025-1411-34, 00025-1411-60, 00025-1411-90, 00025-1412-34, 00025-1412-60, 00025-1415-20, 00025-1415-34, 00025-1415-60, 00025-1416-31, 00025-1416-34, 00025-1416-60, 00172-4430-49, 00172-4430-59, 00172-4431-49, 00172-4431-60, 00247-0416-40, 00247-0416-60, 00440-7829-02, 00440-7829-04, 00440-7829-08, 00591-0397-19, 00591-0397-60, 00591-0398-19, 00591-0398-60, 00904-6806-61, 00904-6807-61, 10135-0745-60, 10135-0745-62, 10135-0746-60, 10544-0949-14, 12280-0288-60, 16590-0024-60, 16590-0432-60, 16590-0564-60, 18837-0012-60, 18837-0290-60, 21695-0425-60, 21695-0456-60, 23490-7228-01, 23490-9246-06, 23629-0052-01, 23629-0052-10, 33358-0247-60, 33358-0248-00, 33358-0248-20, 33358-0248-30, 33358-0248-60, 35356-0467-60, 38779-2778-05, 38779-2778-08, 38779-2778-09, 42291-0232-60, 42291-0232-90, 42291-0233-60, 42367-0110-06, 42367-0111-06, 42549-0679-60, 42571-0133-60, 42571-0134-60, 43063-0707-02, 43063-0707-04, 43063-0707-06, 43063-0707-08, 43063-0707-13, 43063-0707-20, 43386-0160-06, 43386-0160-12, 43386-0161-01, 43386-0161-06, 43393-0020-04, 43393-0022-04, 43393-0023-04, 43393-0024-04, 43393-0200-04, 43393-0200-48, 43393-0200-60, 43393-0203-04, 49999-0589-60, 49999-0987-60, 50268-0555-60, 51552-1378-03, 51927-3870-00, 52372-0656-01, 52372-0656-02, 52959-0353-12, 52959-0353-28, 52959-0353-30, 52959-0353-40, 52959-0353-60, 52959-0354-01, 52959-0354-12, 52959-0354-30, 52959-0354-40, 52959-0525-14, 52959-0525-20, 52959-0525-30, 52959-0525-60, 52959-0531-20, 52959-0531-30, 52959-0531-42, 52959-0531-60, 52959-0531-90, 52959-0692-12, 52959-0692-28, 52959-0692-30, 52959-0692-40, 52959-0692-60, 52959-0693-12, 52959-0693-16, 52959-0693-20, 52959-0693-28, 52959-0693-30, 52959-0693-40, 52959-0693-60, 53002-0564-00, 53002-0564-60, 54124-0440-30, 54569-2765-00, 54569-2765-01, 54569-2765-02, 54569-3426-00, 54569-3426-01, 54569-4579-00, 54569-6097-00, 54868-2176-00, 54868-2176-01, 54868-2176-03, 54868-2176-04, 54868-4164-00, 54868-4164-01, 54868-4165-00, 54868-4165-01, 54868-4766-00, 54868-4766-01, 55045-2548-08, 55045-3047-09, 55175-5105-03, 55175-5150-01, 55175-5150-04, 55289-0248-28, 55289-0406-15, 55289-0406-30, 55289-0640-02, 55289-0640-04, 55289-0640-06, 55289-0640-08, 55289-0640-13, 55289-0698-02, 55289-0698-04, 55289-0698-08, 55289-0698-30, 55700-0271-30, 55887-0543-30, 55887-0543-60, 55887-0543-82, 55887-0543-90, 57866-3172-02, 57866-3908-01, 57866-3909-01, 58016-0190-02, 58016-0347-00, 58016-0347-10, 58016-0347-12, 58016-0347-14, 58016-0347-15, 58016-0347-20, 58016-0347-28, 58016-0347-30, 58016-0347-60, 58016-0348-00, 58016-0348-20, 58016-0498-00, 58016-0498-30, 58016-0498-60, 58016-0498-90, 58016-0577-00, 58016-0577-02, 58016-0577-03, 58016-0577-05, 58016-0577-30, 58016-0577-60, 58016-0577-73, 58016-0577-89, 58016-0577-90, 58016-0766-00, 58016-0766-02, 58016-0766-03, 58016-0766-30, 58016-0766-60, 58016-0766-73, 58016-0766-89, 58016-0766-90, 58016-0783-00, 58016-0783-02, 58016-0783-03, 58016-0783-30, 58016-0783-60, 58016-0783-73, 58016-0783-89, 58016-0783-90, 58016-4504-00, 58016-4504-10, 58016-4504-12, 58016-4504-14, 58016-4504-15, 58016-4504-20, 58016-4504-30, 58016-4504-60, 59762-0028-01, 59762-0028-02, 59762-0028-04, 59762-0029-01, 59762-0029-03, 59762-5007-01, 59762-5007-02, 59762-5008-01, 59762-5008-02, 60687-0735-01, 60687-0735-11, 60687-0746-01, 60687-0746-11, 62991-3178-01, 63370-0159-12, 63629-3243-01, 63629-3359-01, 63629-3359-02, 63629-3359-03, 63704-0008-01, 65162-0436-03, 65162-0436-06, 65162-0438-06, 66267-0428-40, 68001-0231-06, 68001-0232-06, 68084-0040-01, 68084-0040-11, 68084-0041-01, 68084-0041-11, 68084-0978-32, 68084-0978-33, 68115-0381-15, 68115-0381-30, 68115-0381-60, 68115-0560-60, 68258-9135-01, 69339-0123-06, 69339-0124-06, 70954-0443-10, 70954-0443-20, 70954-0444-10, 70954-0444-20, 71205-0865-00, 71205-0865-30, 71205-0865-55, 71205-0865-60, 71205-0865-72, 71205-0865-90, 71205-0866-00, 71205-0866-30, 71205-0866-55, 71205-0866-60, 71205-0866-72, 71205-0866-90, 72789-0111-08, 72789-0121-08, 72789-0128-04, 72789-0128-13, 72789-0300-04, 72789-0300-08, 72789-0300-20, 75834-0264-60, 75834-0265-60, 75839-0159-00, 82804-0037-10), state-specific medication abortion codes from broad cohort inclusion | Day after index mifepristone – 45 days post index mifepristone |
| Subsequent procedure | 59840, 59841, 59812, 59820, 59821, 58120 | Day after index mifepristone – 45 days post index mifepristone |
